# Habit Reversal Training for Tic Disorders: Clinical Outcomes from a Large Sample of Youth and Adults Treated with Therapist-delivered Video Therapy

**DOI:** 10.1101/2025.09.11.25335505

**Authors:** Jamie D. Feusner, Clare C. Beatty, Patrick B. McGrath, Nicholas R. Farrell, Mia Nuñez, Nicholas Lume, Larry Trusky, Stephen M. Smith, Andreas Rhode

## Abstract

Tic disorders, including persistent motor and vocal tic disorders and Tourette syndrome, are neurodevelopmental conditions affecting youth and adults. Habit reversal training (HRT) is a behavioral intervention with demonstrated efficacy from in-person studies, but little information exists about its effectiveness as a remotely-delivered treatment. We examined the effectiveness of therapist-delivered video therapy (HRT) with digital support across the lifespan in a sample of 167 patients with tic disorders (76 children, 38 adolescents, 53 adults). HRT sessions were delivered as one-on-one therapist-delivered video HRT with between-session support including messaging with the therapist, digital support tools, and an online peer community. Tic clinical assessments were administered at baseline, near sessions 7 and 14, and the final treatment session. At session 14, tic symptoms showed a median 39.0% severity reduction (youth: 40.9%; adults: 38.1%). Improvements were maintained through the final treatment sessions, with a median reduction of 44.4% from baseline (youth: 43.5%; adults: 55.4%). This observational analysis demonstrates that therapist-delivered video HRT can effectively reduce tic severity, with maintenance and further improvement of symptoms over an extended treatment duration, in a real-world setting. The virtual delivery format may help address barriers to accessing evidence-based care for tic disorders across the lifespan.

**Strengths and limitations of this study:**

- Large naturalistic sample (N=167) provides real-world effectiveness data for therapist-delivered video HRT across developmental stages from children to adults
- Video therapy format with digital support eliminates geographical barriers while allowing treatment delivery in patients’ natural environments
- Retrospective observational design without control group precludes causal inferences about treatment effectiveness compared to alternative interventions
- Self-report and parent-report outcome measures may introduce reporting bias
- Validated clinical response thresholds are available only for youth measures (PTQ) but not for adult measures (ATQ), limiting interpretation of treatment response rates across age groups

## 1. Introduction

Tic disorders, including persistent (chronic) motor and vocal tic disorders and Tourette syndrome^1^, are common neurodevelopmental conditions characterized by sudden, rapid, recurrent, non-rhythmic motor movements or vocalizations. These disorders affect a substantial portion of the population, with prevalence estimates in children and adolescents ranging from 1.5% to 9.9%^2–4^. Tourette syndrome, specifically, affects approximately.60% to.77% of children, with higher rates observed in boys than girls^5,6^. While likely less common in adults, with prevalence rates ranging from 0.3% to 4.5%^7^, tic disorders can persist throughout the lifespan, with adult-onset cases being relatively rare^8^.

Without treatment, tic disorders can significantly impair quality of life across multiple domains. Youth with chronic tic disorders experience mild to moderate functional impairment in physical, social, familial, academic, and psychological areas, with tic severity often correlating with the level of impairment^9,10^. Similarly, adults report functional limitations affecting social, occupational, and psychological functioning^9^. Both children with tic disorders and their families report lower quality of life compared to those without these conditions^11,12^.

Fortunately, tic disorders can be effectively managed through behavioral therapy. Habit reversal training (HRT), a behavioral intervention that guides patients in increasing their awareness of tics and subsequently using a “competing response” to prevent expression of tics, practicing skills across different settings, and incorporating social supports^13^, has established efficacy for tic disorders. Building upon HRT, Comprehensive Behavioral Intervention for Tics (CBIT) incorporates additional components and has gained recognition as a first-line treatment for tic disorders by major medical academies^14^. HRT remains a foundational behavioral intervention and is similarly recognized as a first-line treatment^15^.Meta-analyses of randomized controlled trials have found HRT to be effective with small to medium effect sizes in adults^16^ and large effect sizes (d=0.80) across age groups^17^. Studies examining HRT implementation in children and adolescents consistently demonstrate significant reductions in tic severity across various delivery formats, including individual face-to-face sessions, group therapy settings, parent-administered home-based treatment, and telehealth approaches^18,19^.

However, HRT and CBIT require specialty-trained therapists and thus are not readily available to everyone with tic disorders because of limited numbers of trained therapists, as well as cost and geographical limitations^20,21^. This shortage of expertise leads to significant challenges in accessing evidence-based care. Furthermore, the absence of specific national guidelines for assessing and treating tic disorders in some regions contributes to inadequate care pathways and increased healthcare utilization and costs^22^.

To address the challenges of delivering evidence-based behavioral therapy in terms of barriers to access, remotely delivered behavioral interventions have shown promising results. Telehealth approaches provide significant practical advantages, such as cost-effectiveness and flexible scheduling, which help reduce the time and financial burden associated with commuting to in-person sessions^23^. The widespread adoption of smartphones among American adults (91-92%; https://www.pewresearch.org/internet/fact-sheet/mobile/) further enhances the feasibility of implementing telehealth approaches, making evidence-based tic disorder treatments more accessible to diverse populations. Remote treatments, in general, have the added benefit of allowing therapy to take place in the patient’s everyday environment, enhancing ecological validity and generalization of skills.

Remotely-delivered behavioral therapy has been studied in several controlled and uncontrolled clinical trials. Videoconference and Voice over Internet Protocol (VoIP) delivery of CBIT has demonstrated efficacy comparable to face-to-face therapy in small randomized controlled trials and pilot studies with both children and adults^23–25^, achieving significant tic reduction with medium-to-large effect sizes (d=0.66-1.31), high treatment satisfaction, and strong therapeutic alliance. Research on remotely-delivered HRT has been more limited. Initial evidence comes from a small pilot study that tested videoconference-delivered HRT in three children ^26^ Another pilot study demonstrated feasibility of a “blended therapy” approach using HRT that combined face-to-face sessions with online coaching^27^. A randomized controlled trial in 40 youth with Tourette syndrome found that online remote behavior therapy offering either HRT or ERP (based on clinical appropriateness) was equally effective as face-to-face delivery of the same interventions^28^.

However, there remain significant gaps in our understanding of HRT effectiveness when delivered remotely. First, while previous studies have examined either basic telehealth delivery or self-guided digital interventions separately, no studies have examined the combination of therapist-delivered video therapy with digital support components (messaging, tracking tools, peer communities). Second, there remains a gap in understanding remote HRT effectiveness in real-world clinical settings across the lifespan, as most research has been conducted in controlled trial environments with limited age ranges. No studies to date have examined the implementation and outcomes of therapist-delivered video HRT with digital support in a large naturalistic sample of both youth and adults with tic disorders. Thus, the objective of this study was to examine treatment outcomes in a large naturalistic sample of individuals with tic disorders who received HRT treatment via an online specialty therapy platform between May 2021 and May 2025. NOCD delivers HRT through therapist-delivered video therapy sessions with supplemental support including therapist messaging, digital tools, and peer community access. Analyzing outcomes across different age groups (children, adolescents, and adults) and examining clinical outcome trajectories over time can provide insights into the real-world utility of comprehensive digital behavioral interventions for tic disorders throughout the lifespan.

## 2. Method

### 2.1 Sample

This was a retrospective, observational longitudinal analysis of clinical data from individuals who received HRT treatment for tic disorders through NOCD’s online specialty therapy platform between May 2021 and May 2025. We included patients who completed a baseline assessment during session 1 or 2, at least one tics assessment at any point after baseline, and a minimum of five tic-focused treatment sessions. The five-session threshold was established based on clinical judgment regarding the minimum number of sessions to reach at least the beginning of core HRT components (awareness training, competing response development, and initial practice) after initial assessments, psychoeducation, and treatment planning. With fewer than five sessions, a patient would typically still be in the preliminary phases and would not have meaningfully “started” the active behavioral treatment components of HRT.

The final analysis included 167 tic disorder patients (qualifying from the inclusion criteria), including children (45.5%), adolescents (22.8%), and adults (31.7%) with primary diagnoses of tic disorders. A detailed patient flow diagram and additional sample characteristics are provided in the Supplementary Materials (Figure S1).

### 2.2 Initial Evaluation and Clinical Assessments

Initial evaluations were conducted by trained therapists using the Diagnostic Interview for Anxiety, Mood, and Obsessive-Compulsive and Related Neuropsychiatric Disorders (DIAMOND^1,29^). Patients who met DSM-5 criteria for persistent (chronic) motor or vocal tic disorder, or Tourette syndrome, were eligible for treatment. Those rated as having “extreme” symptom severity were typically referred to more intensive treatment options (see Supplementary Material).

### 2.3 Treatment Approach

HRT was delivered through structured therapist-delivered video therapy. Therapists used a secure version of Zoom that was US HIPAA (Health Insurance Portability and Accountability Act)-compliant and compliant with other countries’ health information privacy regulations. Patients used personal computing or mobile devices, and had live technical support available during business hours to address connectivity issues. Treatment included weekly 60-minute video sessions and involved core HRT components (awareness training, competing response training, generalization training, and social supports^13^) and incorporated stimulus control strategies and functional analysis of the tics. As a complement to HRT treatment, patients also had access to other elements of care that are available to all NOCD Members. This included access to support groups, a moderated online community, and peer support individuals called Member Advocates. The participants also had access to application tools including the ability to message one’s therapist, view one’s symptom reduction data graphically, and engage in symptom tracking. These elements were optional. Treatment was delivered by licensed mental health professionals who held a minimum of a master’s or doctoral degree in psychology, social work, counseling, or related mental health fields. All therapists were required to maintain appropriate state licensure. In addition to their formal education, therapists completed an intensive 12-week training program specifically in OCD-related disorders and HRT, followed by ongoing clinical supervision and consultation.

### 2.4 Assessments

The primary outcome measures were the Parent Tic Questionnaire (PTQ) and Adult Tic Questionnaire (ATQ), administered in parent-report and self-report formats, respectively. These questionnaires assess the frequency and intensity of specific motor and vocal tics, with up to 14 motor tics and 13-14 vocal tics evaluated. For each tic present, respondents rate both frequency and intensity on a 1-4 scale. Frequency ratings range from 1 (weekly/few times) to 4 (constantly), while intensity ratings range from 1 (very mild) to 4 (extremely forceful). A severity score (2-8) is calculated for each tic by summing its frequency and intensity ratings. Motor and vocal tic scores range from 0-112 each, with a total tic score ranging from 0-224. Both questionnaires demonstrate excellent internal consistency (PTQ: α=.80-.86; ATQ: α=.86-.91) and test-retest reliability (PTQ: ICC=.84-.89; ATQ: r=.81-.88), with strong convergent validity with clinician-rated measures like the Yale Global Tic Severity Scale (r=.62-.73) and good discriminant validity from measures of ADHD, OCD, and externalizing behaviors^30,31^.

Secondary measures assessed depression, anxiety, and stress ((Depression, Anxiety, and Stress Scale; [DASS-21^32^] and DASS-Youth [DASS-Y^33^), quality of life (Quality of Life Enjoyment and Satisfaction Questionnaire-Short Form [Q-LES-Q-SF^34^] for adults and Pediatric Quality of Life Enjoyment and Satisfaction Questionnaire [PQ-LES-Q^35^] for youth and disability and functioning (World Health Organization Disability Assessment Schedule 2.0 [WHODAS 2.0^36^] in adults). See Supplementary Material for complete measure descriptions and youth results.

### 2.5 Data Processing

We identified tic-focused treatment sessions using ICD-10 diagnostic codes (F95.0-F95.9) from clinical records. Due to the naturalistic clinical setting, baseline assessments were not always completed during the first session. Therefore, we implemented a flexible baseline approach that defined baseline as the first tic assessment completed during either session 1 or 2. We analyzed outcomes at sessions 7 and 14 to capture both mid-treatment effects (consistent with typical 8-10 session protocols) and extended treatment outcomes (consistent with 12-16 session protocols and booster session approaches)^37^. When assessments were missed, we used a last-observation-carried-forward (LOCF) approach, where a patient’s most recent assessment was used until a new assessment was completed. This conservative method ensures we did not overestimate treatment effects when data were missing.

### 2.6 Data Analysis

Treatment parameters were calculated based on each patient’s session attendance patterns, including total sessions completed and time (weeks) between sessions. Treatment engagement metrics (messaging, app opens, session attendance) were analyzed to examine platform usage patterns.

Primary and secondary outcomes were analyzed using linear mixed models with treatment session (baseline, session 7, session 14) as a fixed factor, patient as a random factor, and respective outcome scores (tic severity, depression, anxiety, stress, quality of life, and disability) as dependent variables. Analyses were conducted separately for youth and adult samples to examine age-specific treatment effects. We also examined outcomes at each patient’s final treatment session (regardless of session number). Linear mixed models included all patients in the analyzed sample regardless of missing data at post-baseline timepoints, consistent with intent-to-treat principles. For calculations requiring paired observations (effect sizes, percent reductions, and response rates), analyses were limited to patients with data at both the baseline and comparison timepoints. For all primary and secondary outcomes, absolute point changes and percentage changes were calculated. Percentage changes were calculated at the individual patient level and then aggregated to determine mean and median percent reductions.

Individual-level changes were calculated by comparing baseline scores with assessments at session 7, session 14, and each patient’s final treatment session. Treatment response categories were based on uniform percentage thresholds applied consistently across all age groups: ≥55% reduction, ≥45% reduction, ≥35% reduction, ≥25% reduction, >0% reduction (any improvement), and worsening.

Response rates were examined for the overall sample and stratified by age group (children: 5-12, adolescents: 13-17, adults: 18+). For individual-level effects, median percent improvements with interquartile ranges and response rates across clinically meaningful thresholds are reported. Effect sizes were calculated using Hedges’ g with a paired-samples approach, which divides the mean difference by the standard deviation of the differences rather than the pooled standard deviation of raw scores. This method accounts for within-subject correlation by controlling for each patient’s baseline variability, producing more conservative but methodologically appropriate estimates^38^.

Reliable Change Index (RCI) analyses were conducted to identify statistically reliable individual-level changes that exceed measurement error^39^. The RCI was calculated using Jacobson and Truax’s (1991) method, based on published test-retest reliability coefficients (PTQ ICC=.89; ATQ r=.87) and baseline standard deviations (PTQ=20.55; ATQ=25.00)^31,40^. For the PTQ (youth), a reduction of ≥18.89 points was required to meet the threshold for reliable change; for the ATQ (adults), a reduction of ≥24.99 points was needed. Statistically derived RCI classifications were compared to the uniform percentage-based response thresholds (≥55% reduction, ≥45% reduction, ≥35% reduction, ≥25% reduction) to assess concordance between statistical and clinical significance RCI classifications were examined at both session 14 and the final treatment session.

Factors potentially influencing treatment response were examined. Baseline symptom severity and treatment duration were tested as potential moderators of treatment effects using linear mixed models with interaction terms (timepoint × moderator × age group). These analyses assessed whether these factors affected treatment response differently in youth versus adults. Pearson correlations were calculated between each moderator (baseline severity, treatment duration) and both absolute and percent reduction in tic symptoms at sessions 7 and 14. Exploratory analyses also examined the impact of baseline severity on treatment outcomes. Additionally, for patients who had ≥14 sessions (n=92), we analyzed continued improvement by comparing session 14 scores to final treatment session scores using linear mixed models to assess whether patients demonstrated significant additional therapeutic gains beyond the primary treatment phase of 14 sessions.

Analyses were conducted separately for adult and youth participants using age-appropriate measures. Adult analyses (n=53) included demographics, baseline tic severity, and adult-specific measures (DASS-21, WHODAS-2.0, Q-LES-Q-SF). Youth analyses (n=114) included demographics, baseline tic severity, and youth-specific measures (DASS-Y, PQ-LES-Q). Analyses were conducted using R Statistical Software, with additional methodological details available in the Supplementary Material.

### 2.7 Ethical Considerations

The analyses in this study did not require research ethics board review as this does not meet the criteria for Human Subject Research as defined by federal regulations for human subject protections, 45 CFR 46.102(e); namely, this is a secondary analysis of de-identified data from clinical records, obtained and analyzed retrospectively, and was not the result of a research intervention or interaction. The UCLA IRB Office confirmed that this study did not meet the criteria for Human Subject Research (PRE#20-008583) and thus did not require approval. Compliance with data protection laws was ensured through NOCD’s privacy policy, which all patients agreed to, outlining data use and protection measures. This retrospective analysis was not prospectively registered as it involved secondary analysis of existing clinical data.

### 2.8 Patient and Public Involvement

Patients and public were not involved in setting the research agenda, study design, or conduct of this research as this was a retrospective analysis of de-identified clinical data from routine care delivery. No patients were involved in the interpretation of results or preparation of the manuscript.

## 3. Results

### 3.1 Sample

The initial sample consisted of 387 patients treated for tics in the analysis period. After applying the analysis inclusion/exclusion criteria, 220 patients (56.8%) were excluded for the following reasons: no baseline assessment in sessions 1-2 (135 patients, 61.4% of exclusions), no post-baseline assessments (65 patients, 29.5% of exclusions), and fewer than 5 tic-focused sessions (20 patients, 9.1% of exclusions).

The final analysis included 167 patients with tic disorders. The sample showed a predominance of youth patients, with 68.3% under age 18 and 31.7% adult. The youth sample was divided between children (45.5% of total; mean age=9.8 years, SD=1.7) and adolescents (22.8% of total; mean age=14.7 years, SD=1.4). Adults comprised 31.7% of the sample (mean age=34.8 years, SD=12.4). Overall, the mean age for the entire sample was 18.9 years (SD=13.1) (Table 1). Missing data analyses indicated no significant differences between early discontinuation and sustained engagement groups for age, baseline tic severity, or clinical measures, supporting the Missing Completely At Random assumption (see Supplementary Materials for detailed MCAR analyses).

**Table 1.**
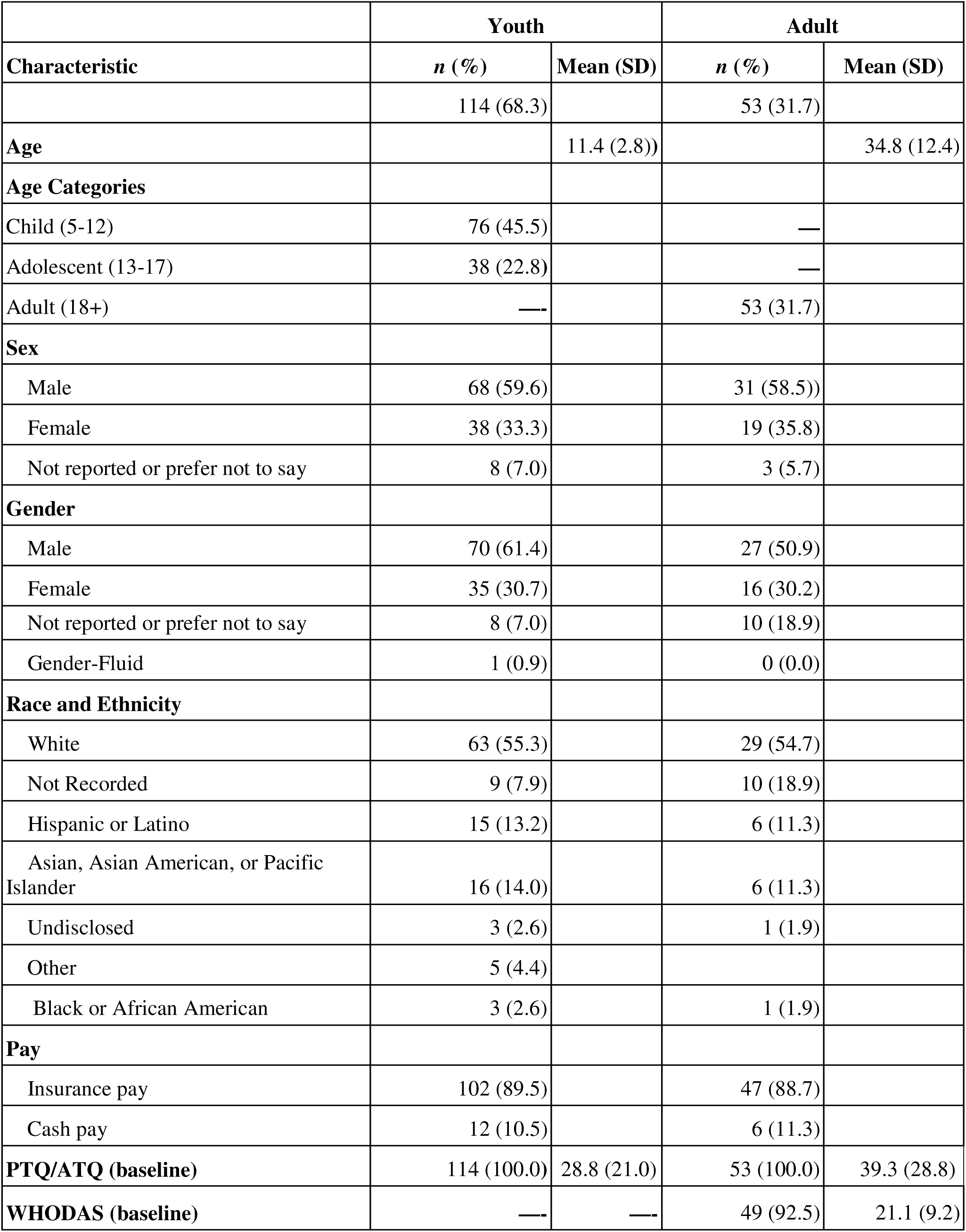
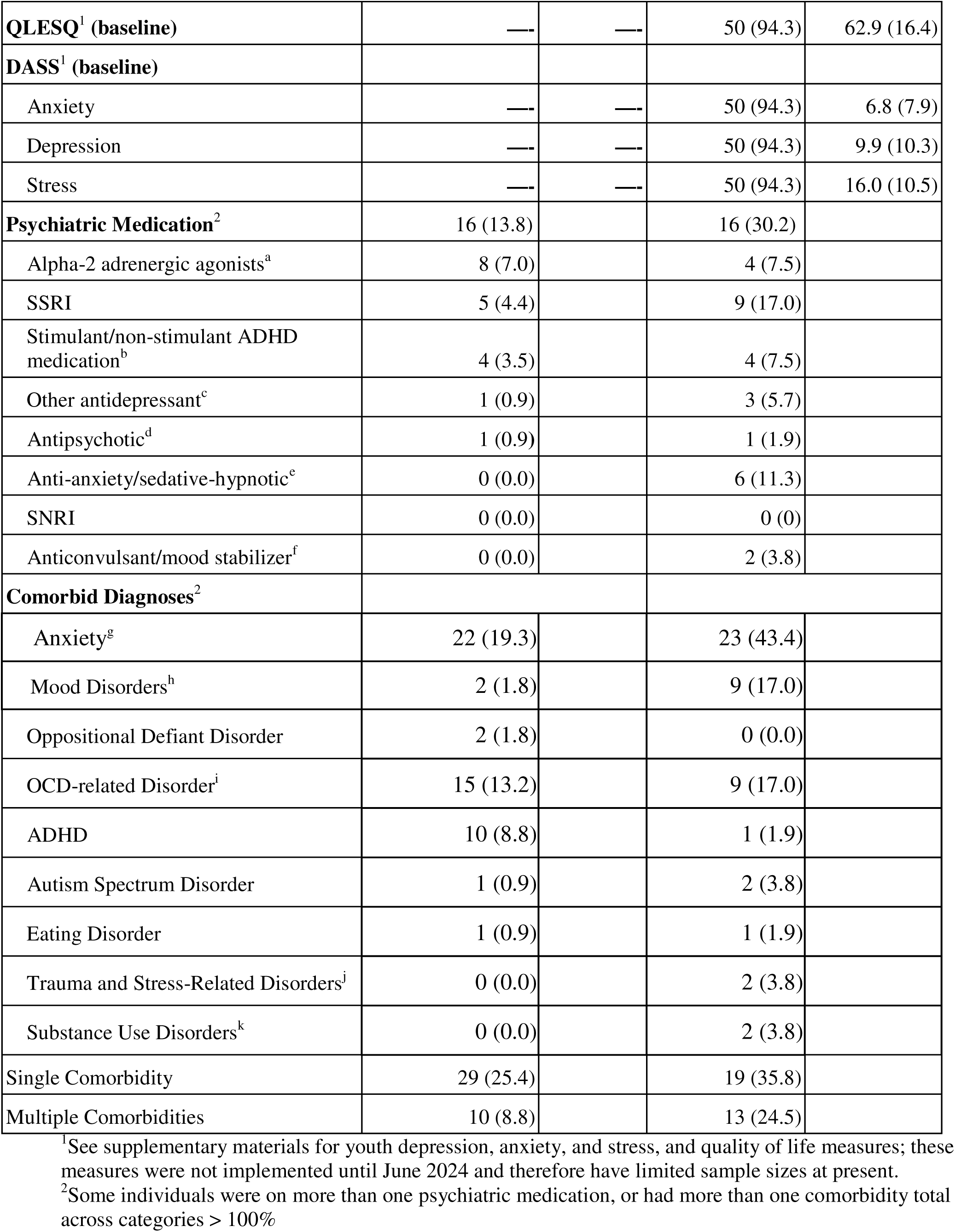
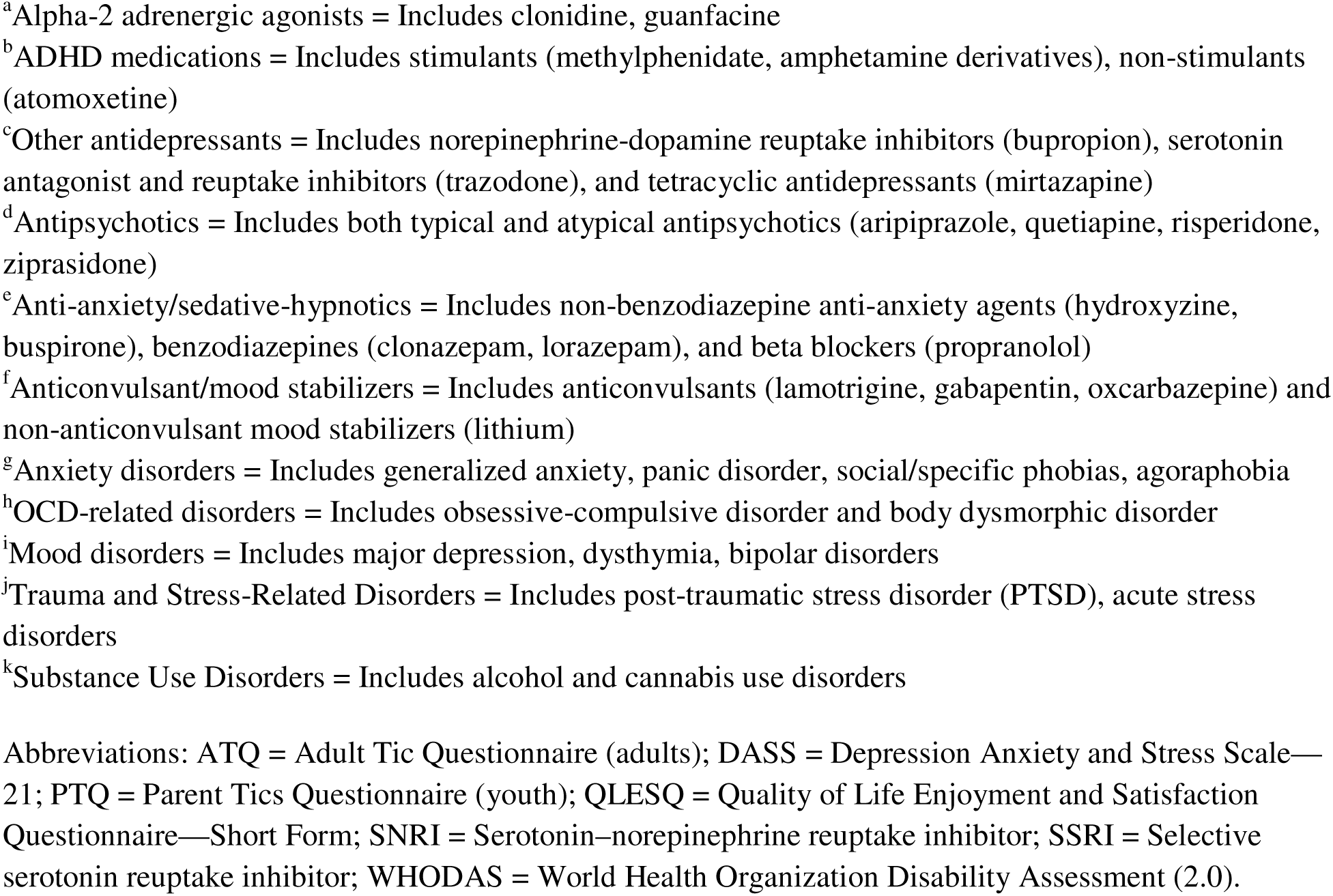
Demographics and psychometrics.

|  | **Youth** |  | **Adult** |  |
| --- | --- | --- | --- | --- |
| **Characteristic** | ***n* (%)** | **Mean (SD)** | ***n* (%)** | **Mean (SD)** |
|  | 114 (68.3) |  | 53 (31.7) |  |
| **Age** |  | 11.4 (2.8)) |  | 34.8 (12.4) |
| **Age Categories** |  |  |  |  |
| Child (5-12) | 76 (45.5) |  | — |  |
| Adolescent (13-17) | 38 (22.8) |  | — |  |
| Adult (18+) | — |  | 53 (31.7) |  |
| **Sex** |  |  |  |  |
| Male | 68 (59.6) |  | 31 (58.5)) |  |
| Female | 38 (33.3) |  | 19 (35.8) |  |
| Not reported or prefer not to say | 8 (7.0) |  | 3 (5.7) |  |
| **Gender** |  |  |  |  |
| Male | 70 (61.4) |  | 27 (50.9) |  |
| Female | 35 (30.7) |  | 16 (30.2) |  |
| Not reported or prefer not to say | 8 (7.0) |  | 10 (18.9) |  |
| Gender-Fluid | 1 (0.9) |  | 0 (0.0) |  |
| **Race and Ethnicity** |  |  |  |  |
| White | 63 (55.3) |  | 29 (54.7) |  |
| Not Recorded | 9 (7.9) |  | 10 (18.9) |  |
| Hispanic or Latino | 15 (13.2) |  | 6 (11.3) |  |
| Asian, Asian American, or Pacific Islander | 16 (14.0) |  | 6 (11.3) |  |
| Undisclosed | 3 (2.6) |  | 1 (1.9) |  |
| Other | 5 (4.4) |  |  |  |
| Black or African American | 3 (2.6) |  | 1 (1.9) |  |
| **Pay** |  |  |  |  |
| Insurance pay | 102 (89.5) |  | 47 (88.7) |  |
| Cash pay | 12 (10.5) |  | 6 (11.3) |  |
| **PTQ/ATQ (baseline)** | 114 (100.0) | 28.8 (21.0) | 53 (100.0) | 39.3 (28.8) |
| **WHODAS (baseline)** | — | — | 49 (92.5) | 21.1 (9.2) |
| **QLESQ<sup>1</sup> (baseline)** | — | — | 50 (94.3) | 62.9 (16.4) |
| **DASS<sup>1</sup> (baseline)** |  |  |  |  |
| Anxiety | — | — | 50 (94.3) | 6.8 (7.9) |
| Depression | — | — | 50 (94.3) | 9.9 (10.3) |
| Stress | — | — | 50 (94.3) | 16.0 (10.5) |
| **Psychiatric Medication<sup>2</sup>** | 16 (13.8) |  | 16 (30.2) |  |
| Alpha-2 adrenergic agonists<sup>a</sup> | 8 (7.0) |  | 4 (7.5) |  |
| SSRI | 5 (4.4) |  | 9 (17.0) |  |
| Stimulant/non-stimulant ADHD medication<sup>b</sup> | 4 (3.5) |  | 4 (7.5) |  |
| Other antidepressant<sup>c</sup> | 1 (0.9) |  | 3 (5.7) |  |
| Antipsychotic<sup>d</sup> | 1 (0.9) |  | 1 (1.9) |  |
| Anti-anxiety/sedative-hypnotic<sup>e</sup> | 0 (0.0) |  | 6 (11.3) |  |
| SNRI | 0 (0.0) |  | 0 (0) |  |
| Anticonvulsant/mood stabilizer<sup>f</sup> | 0 (0.0) |  | 2 (3.8) |  |
| **Comorbid Diagnoses<sup>2</sup>** |  |  |  |  |
| Anxiety<sup>g</sup> | 22 (19.3) |  | 23 (43.4) |  |
| Mood Disorders<sup>h</sup> | 2 (1.8) |  | 9 (17.0) |  |
| Oppositional Defiant Disorder | 2 (1.8) |  | 0 (0.0) |  |
| OCD-related Disorder<sup>i</sup> | 15 (13.2) |  | 9 (17.0) |  |
| ADHD | 10 (8.8) |  | 1 (1.9) |  |
| Autism Spectrum Disorder | 1 (0.9) |  | 2 (3.8) |  |
| Eating Disorder | 1 (0.9) |  | 1 (1.9) |  |
| Trauma and Stress-Related Disorders<sup>j</sup> | 0 (0.0) |  | 2 (3.8) |  |
| Substance Use Disorders<sup>k</sup> | 0 (0.0) |  | 2 (3.8) |  |
| Single Comorbidity | 29 (25.4) |  | 19 (35.8) |  |
| Multiple Comorbidities | 10 (8.8) |  | 13 (24.5) |  |

<sup>1</sup>See supplementary materials for youth depression, anxiety, and stress, and quality of life measures; these measures were not implemented until June 2024 and therefore have limited sample sizes at present.
<sup>2</sup>Some individuals were on more than one psychiatric medication, or had more than one comorbidity total across categories > 100%
<sup>a</sup>Alpha-2 adrenergic agonists = Includes clonidine, guanfacine
<sup>b</sup>ADHD medications = Includes stimulants (methylphenidate, amphetamine derivatives), non-stimulants (atomoxetine)
<sup>c</sup>Other antidepressants = Includes norepinephrine-dopamine reuptake inhibitors (bupropion), serotonin antagonist and reuptake inhibitors (trazodone), and tetracyclic antidepressants (mirtazapine)
<sup>d</sup>Antipsychotics = Includes both typical and atypical antipsychotics (aripiprazole, quetiapine, risperidone, ziprasidone)
<sup>e</sup>Anti-anxiety/sedative-hypnotics = Includes non-benzodiazepine anti-anxiety agents (hydroxyzine, buspirone), benzodiazepines (clonazepam, lorazepam), and beta blockers (propranolol)
<sup>f</sup>Anticonvulsant/mood stabilizers = Includes anticonvulsants (lamotrigine, gabapentin, oxcarbazepine) and non-anticonvulsant mood stabilizers (lithium)
<sup>g</sup>Anxiety disorders = Includes generalized anxiety, panic disorder, social/specific phobias, agoraphobia
<sup>h</sup>OCD-related disorders = Includes obsessive-compulsive disorder and body dysmorphic disorder
<sup>i</sup>Mood disorders = Includes major depression, dysthymia, bipolar disorders
<sup>j</sup>Trauma and Stress-Related Disorders = Includes post-traumatic stress disorder (PTSD), acute stress disorders
<sup>k</sup>Substance Use Disorders = Includes alcohol and cannabis use disorders
Abbreviations: ATQ = Adult Tic Questionnaire (adults); DASS = Depression Anxiety and Stress Scale—21; PTQ = Parent Tics Questionnaire (youth); QLESQ = Quality of Life Enjoyment and Satisfaction Questionnaire—Short Form; SNRI = Serotonin–norepinephrine reuptake inhibitor; SSRI = Selective serotonin reuptake inhibitor; WHODAS = World Health Organization Disability Assessment (2.0).

### 3.2 Treatment Delivery and Engagement

Patients engaged in therapist-delivered video HRT sessions with a median frequency of 0.7 sessions per week [IQR: 0.5-0.9] (approximately 2-3 sessions per month). Overall, patients completed a median of 15 sessions [IQR: 9-24.5] over a median duration of 30.9 weeks [IQR: 14.4-54.9], with no significant differences between youth and adults in total treatment duration (p=0.60). For the primary treatment phase (sessions 1-14), 55.1% of patients (92/167) completed ≥14 sessions over a median duration of 20.9 weeks [IQR: 14.9-27.9]. Among patients who completed ≥14 sessions, 92.4% (85/92) engaged in additional maintenance therapy, completing a median total of 24 sessions over an additional 21.9 weeks. Detailed treatment completion patterns and age-group comparisons are provided in the Supplementary Materials.

### 3.3 Overall Group Mean Treatment Effects

In the combined group sample (F2,239.08=30.92, P<.001), tic severity scores decreased from a mean of 32.1 (SD=24.2) to a mean of 24.3 (SD=19.7) at session 7 (-7.8 points, 24.3%; Hedges g=0.33, 95% CI [0.22, 0.44], n=146), and to a mean of 22.4 (SD=23.3) at session 14 (-8.8 points, 30.2%; Hedges g=0.47, 95% CI [0.31, 0.64], n=92) (see Table 2, Figure 1). At the final session, overall scores decreased further to a mean of 19.1 (SD=20.6), showing a 40.5% reduction from baseline (−13.0 points; Hedges’ *g*=0.57, 95% CI [0.44, 0.70], *n*=167).

**Figure 1.**
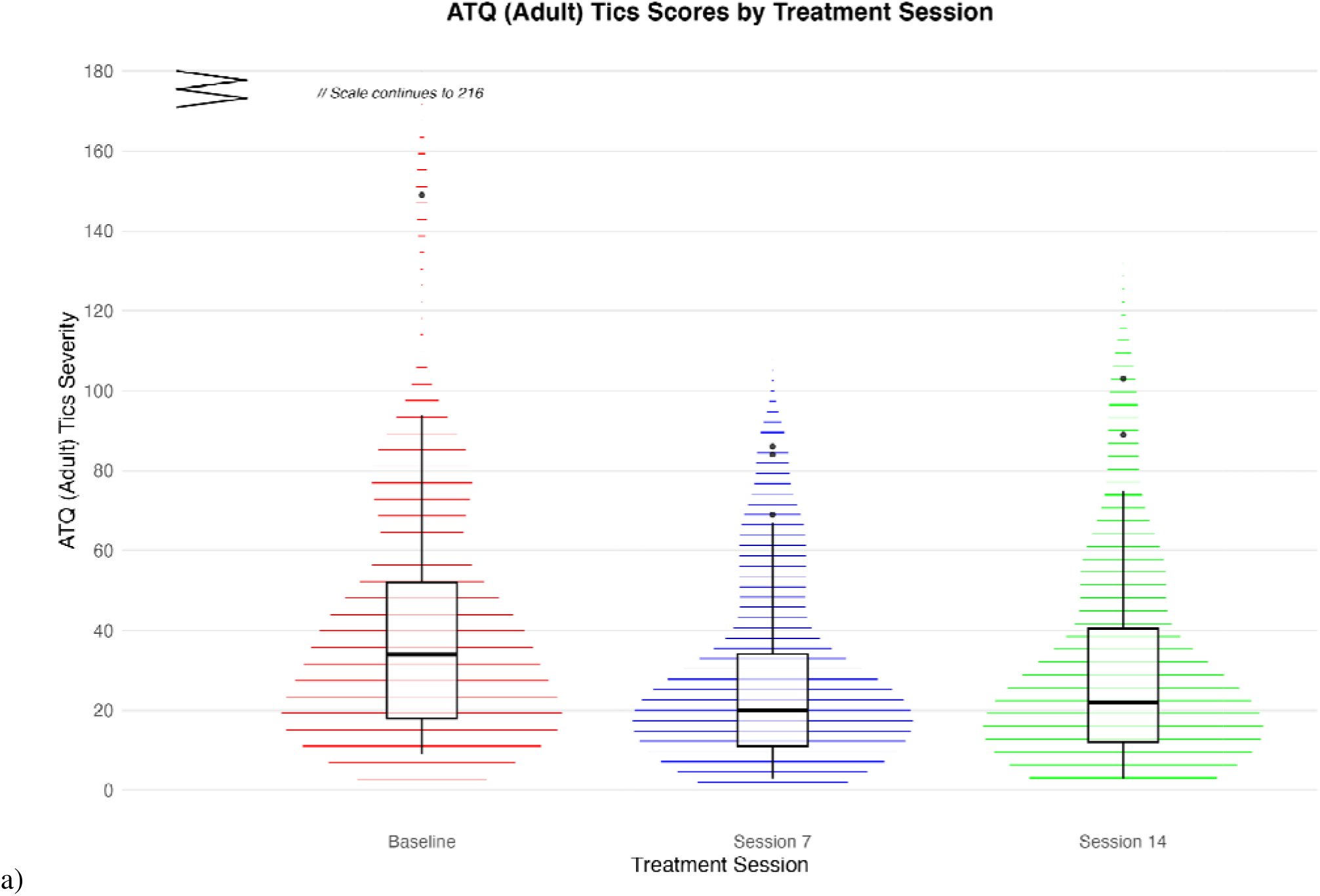
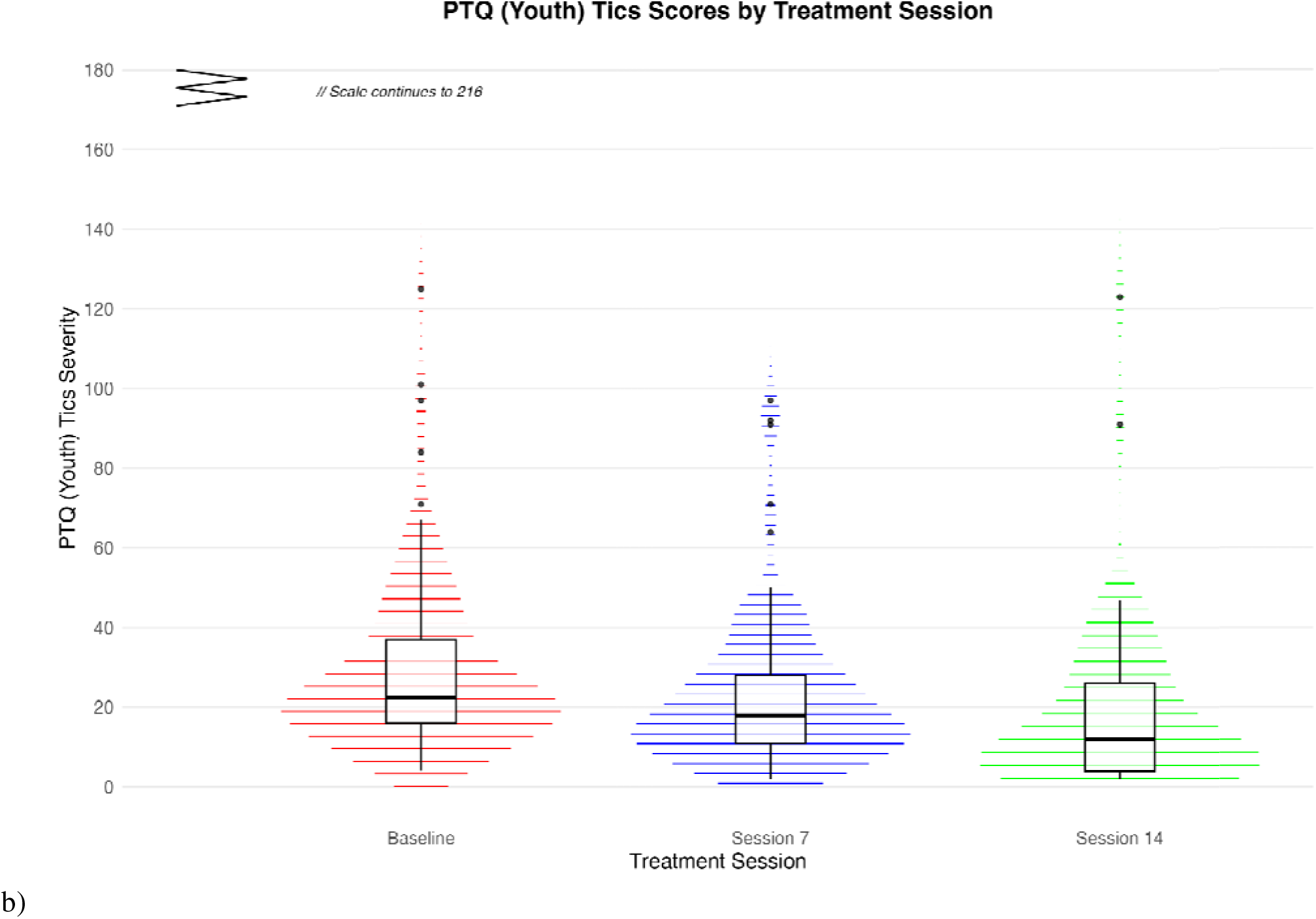
PTQ and ATQ Scores by Session Changes in a) youth (top) and b) adult (bottom) symptoms as assessed by the PTQ and ATQ, respectively, with treatment. Median and interquartile ranges are indicated in the box-and-whisker plots. P <.001 for the effect of assessment period. Abbreviations: ATQ = Adult Tic Questionnaire, PTQ = Parent Tic Questionnaire.

**Table 2.** PTQ and ATQ Scores by Age Group, Condition, and Time Point.

| Age Group |  | Baseline | Session 7 | Session 14 |
| --- | --- | --- | --- | --- |
| Youth | n (%) | 114 (100.0) | 101 (88.6) | 61 (53.5) |
|  | M (SD) | 28.8 (21.0) | 22.6 (18.5) | 18.6 (21.3) |
| Adults | n (%) | 53 (100.0) | 45 (84.9) | 31 (58.8) |
|  | M (SD) | 39.3 (28.8) | 28.0 (22.0) | 29.8 (25.7) |

Abbreviations: ATQ = Adult Tic Questionnaire, M = Mean, PTQ = Parent Tic Questionnaire, SD = Standard Deviation

The youth group showed significant improvement in tic symptoms (F2,168.88=18.57, P<.001), with PTQ scores decreasing from a mean of 28.8 (SD=21.0, n=114) to a mean of 22.7 (SD=18.5) at session 7 (-6.1 points, 21.2%; Hedges g=0.30, 95% CI [0.17, 0.43], n=101), and to a mean of 18.6 (SD=21.3) at session 14 (-10.2 points, 35.4%; Hedges g=0.53, 95% CI [0.30, 0.77], n=61) (see Figure 1a). At the final session, PTQ scores further decreased to a mean of 16.8 (SD=16.2), reflecting a 41.7% improvement from baseline (−12.0 points; Hedges’ *g*=0.63, 95% CI [0.43, 0.82], *n*=114) (see Figure 2a).

**Figure 2.**
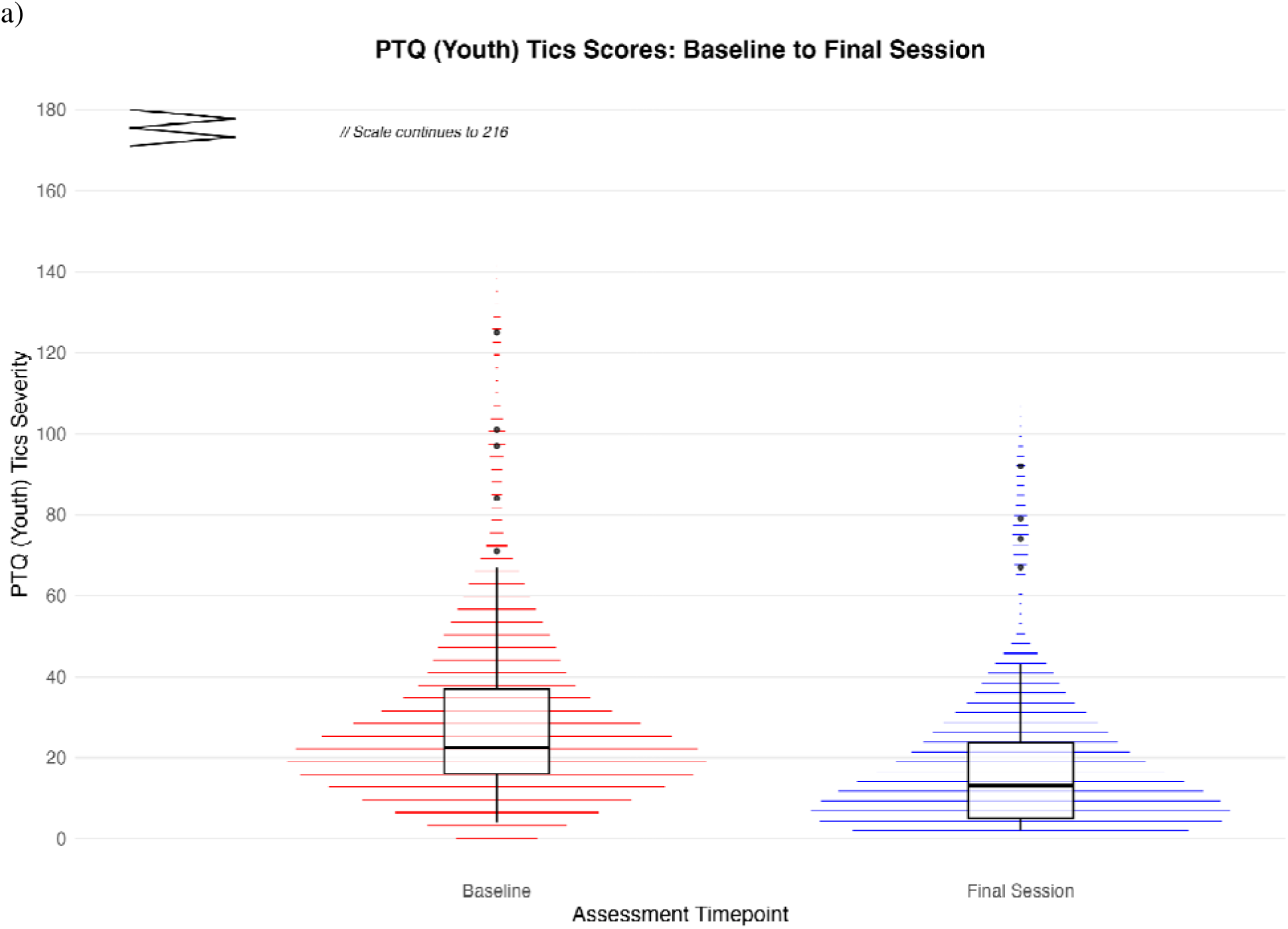
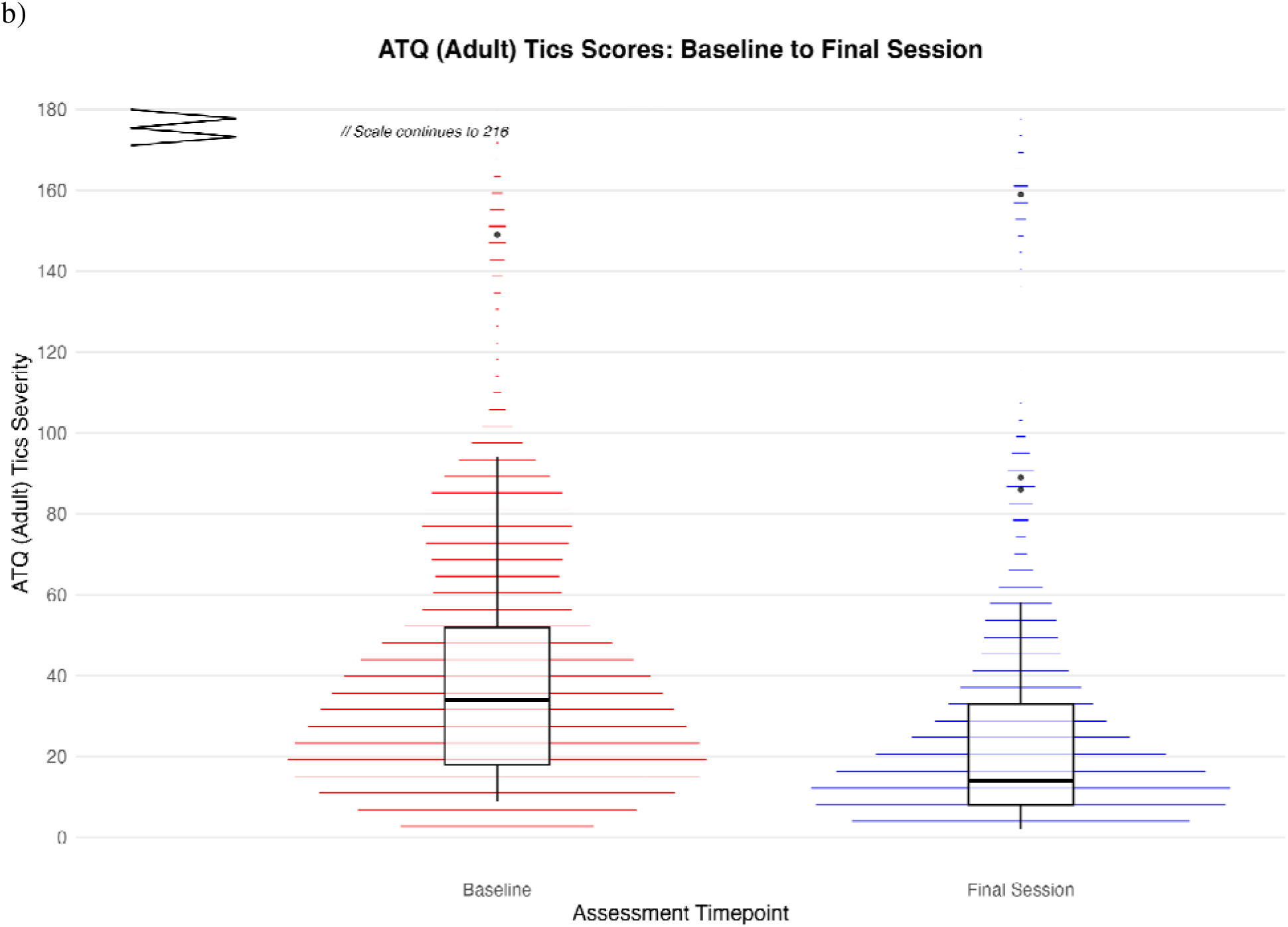
Final Session PTQ and ATQ Scores Changes in a) youth (N=114; top) and b) adult (N=53; bottom) tic severity from baseline to final session (range: 5-72 sessions, median: 15 sessions). Median and interquartile ranges are indicated in the box-and-whisker plots. P <.001 for the effect of assessment period. Abbreviations: ATQ = Adult Tic Questionnaire, PTQ = Parent Tic Questionnaire.

The adult group also demonstrated significant improvement in tic symptoms (F2,73.44=15.08, P<.001), with ATQ scores decreasing from a mean of 39.3 (SD=28.8) to a mean of 28.0 (SD=22.0) at session 7 (-11.3 points, 28.8%; Hedges g=0.40, 95% CI [0.21, 0.59], n=45), and to a mean of 29.8

(SD=25.7) at session 14 (-9.5 points, 24.2%; Hedges g=0.36, 95% CI [0.17, 0.54], n=31) (see Figure 1b). At the final session, ATQ scores further decreased to a mean of 24.2 (SD=27.4), reflecting a 38.4% improvement (−15.1 points; Hedges’ *g*=0.53, 95% CI [0.35, 0.71], *n*=53) (see Figure 2b).

#### 3.3.1 Continued improvement beyond session 14

Among the 92 patients who completed ≥14 sessions, 92.4% (85/92) engaged in at least one maintenance therapy session, completing a median of 10 additional sessions [IQR: 3-19] over a median duration of 21.9 weeks [IQR: 7.9-42.9]. For these 85 patients, significant continued improvement was observed from session 14 to final session (F[,[[=11.38, P=0.001). Overall tic severity scores decreased an additional 4.9 points from session 14 (mean=22.3, SD=22.8) to final session (mean=17.4, SD=16.2), representing a small but significant effect (Hedges’ g=0.22, 95% CI [0.09, 0.35]). The youth group (n=57) showed continued improvement with PTQ scores decreasing an additional 3.9 points from session 14 (mean=19.2, SD=21.8) to final session (mean=15.4, SD=15.6; F[,[[=4.94, P=0.030; Hedges’ g=0.18, 95% CI [0.02, 0.35]). The adult group (n=28) demonstrated the largest continued improvement, with ATQ scores decreasing an additional 6.9 points from session 14 (mean=28.4, SD=24.0) to final session (mean=21.6, SD=16.8; F[,[[=7.19, P=0.012; Hedges’ g=0.28, 95% CI [0.07, 0.50]).

#### 3.3.2 Baseline severity effects

A three-way interaction model (timepoint × baseline severity × age group) revealed that baseline severity influences treatment trajectories differently between youth and adults F(2, 297.13)=6.74, p=.001. Among youth, baseline severity was positively associated with absolute symptom reduction at both session 7 (r(99)=.50, p <.001) and session 14 (r(59)=.51, p <.001), but not with percent reduction at either timepoint (session 7: r(99)=.18, p=.08; session 14: r(59)=.08, p=.53). This indicates that youth with higher initial severity achieve larger point reductions while maintaining similar relative improvement rates as those with milder symptoms. Adults showed a different pattern. At session 7, baseline severity was associated with greater absolute reduction (r(43)=.47, p=.001) but not percent reduction (r(43)=.03, p=.86). By session 14, this relationship disappeared for absolute reduction (r(29)=–.03, p=.88) and trended negative for percent reduction (r(29)=–.30, p=.10). These findings indicate that baseline severity influences treatment response patterns differently across age groups, with youth showing consistent relationships between initial severity and absolute improvement throughout treatment, while adults demonstrate more variable patterns that change over the course of therapy.

*Treatment duration effects.* Additional analyses revealed no significant relationship between treatment duration and symptom improvement in either age group (see Supplementary Materials for detailed analyses).

### 3.4 Individual-Level Changes and Response Rates

For youth (n=114), median percent improvement was 24.1% [IQR:-2.3–50.0%] at session 7 and 40.9% [IQR: 12.5–76.5%] at session 14. At session 14, 42.6% (26/61) achieved ≥55% reduction, 49.2% (30/61) had ≥45% reduction, 55.7% (34/61) had ≥35% reduction, 68.9% (42/61) had ≥25% reduction, 77.0% (47/61) had >0% reduction, and 18.0% (11/61) showed worsening. At final session, median percent improvement in tic severity was 43.5% [IQR: 12.9–72.0%], with 36.8% (42/114) achieving ≥55% reduction, 46.5% (53/114) achieving ≥45% reduction, 62.3% (71/114) achieving ≥35% reduction, 70.2% (80/114) achieving ≥25% reduction, 78.9% (90/114) had >0% reduction, and 18.4% (21/114) showed worsening.

For adults (n=53), median improvement was 19.0% [IQR: 5.0–50.0%] at session 7 and 38.1% [IQR: 8.7–51.6%] at session 14. At session 14, 22.6% (7/31) achieved ≥55% reduction, 29.0% (9/31) had ≥45% reduction, 51.6% (16/31) had ≥35% reduction, 61.3% (19/31) had ≥25% reduction, 80.6% (25/31) had >0% reduction, and 19.4% (6/31) showed worsening. At final session, median improvement was 55.4% [IQR: 19.0–64.6%], with 50.9% (27/53) achieving ≥55% reduction, 56.6% (30/53) achieving ≥45% reduction, 66.0% (35/53) achieving ≥35% reduction, 69.8% (37/53) achieving ≥25% reduction, 86.8% (46/53) had >0% reduction, and 13.2% (7/53) showed worsening.

In the combined sample (n=167), median improvement in tic severity was 39.0% [IQR: 9.4– 71.4%] at session 14 and 44.4% [IQR: 16.7–69.8%] at final session. At session 14, 54.3% (50/92) achieved ≥35% reduction and 35.9% (33/92) achieved ≥55% reduction. At final session, 63.5% (106/167) achieved ≥35% reduction and 41.3% (69/167) achieved ≥55% reduction. No statistically significant differences were found between age groups at any timepoint (all p>0.05).

### 3.5 Reliable Change and Concordance with Clinical Response

At session 14, 21.3% of youth (n=61) and 9.7% of adults (n=31) met criteria for reliable improvement. At the final session, 22.8% of youth (n=114) and 22.6% of adults (n=53) demonstrated reliable change. Concordance between RCI-defined improvement and percentage-based response thresholds varied by threshold level. Agreement was highest for ≥55% reduction (77.2% meeting RCI-defined improvement at session 14, 74.3% at final session) and lower for ≥35% reduction (60.9% at session 14, 58.1% at final session).

### 3.6 Depression, Anxiety, and Stress, Quality of Life and Disability

Adults demonstrated consistent improvement across several secondary symptom domains at session 14 (see Table 3). Significant reductions were observed in stress (median reduction 12.5%, IQR: - 15.4–53.3%; g=0.33, 95% CI [-0.03, 0.69], p<.001), with smaller improvements in anxiety (median reduction 0.0%, IQR:-58.3–63.3%; g=0.14, 95% CI [-0.20, 0.48], p=.074) and depression (median reduction 0.0%, IQR:-100.0–60.7%; g=0.01, 95% CI [-0.33, 0.35], p=.316). Quality of life improved modestly (median increase 0.7%, IQR:-11.3–14.9%; g=0.03, 95% CI [-0.22, 0.27]), and functional impairment decreased (median reduction 0.0%, IQR: 0.0–13.9%; g=0.23, 95% CI [0.02, 0.44], p<.001). At the final session adults showed enhanced improvement patterns with larger effect sizes across most domains: stress (g=0.67, 95% CI [0.33, 1.02]), anxiety (g=0.34, 95% CI [0.07, 0.61]), depression (g=0.22, 95% CI [-0.02, 0.46]), quality of life (g=0.22, 95% CI [0.00, 0.45]), and functional impairment (g=0.35, 95% CI [0.11, 0.60]), suggesting continued benefits with extended treatment duration. Youth depression, anxiety, and stress, and quality of life measures were not implemented until June 2024 and therefore have limited sample sizes (n=14–15); see Supplementary Materials for youth results.

**Table 3.**
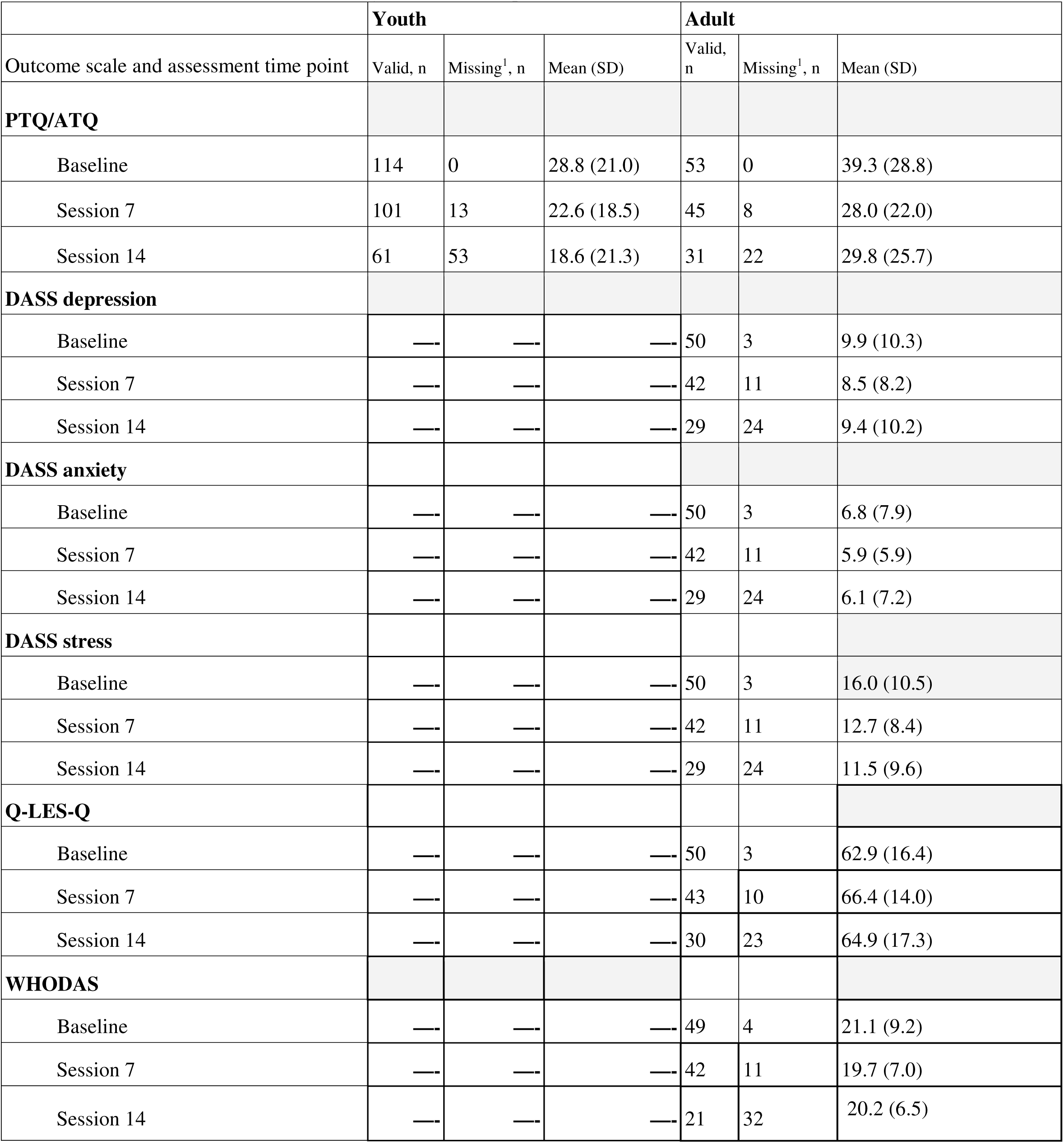
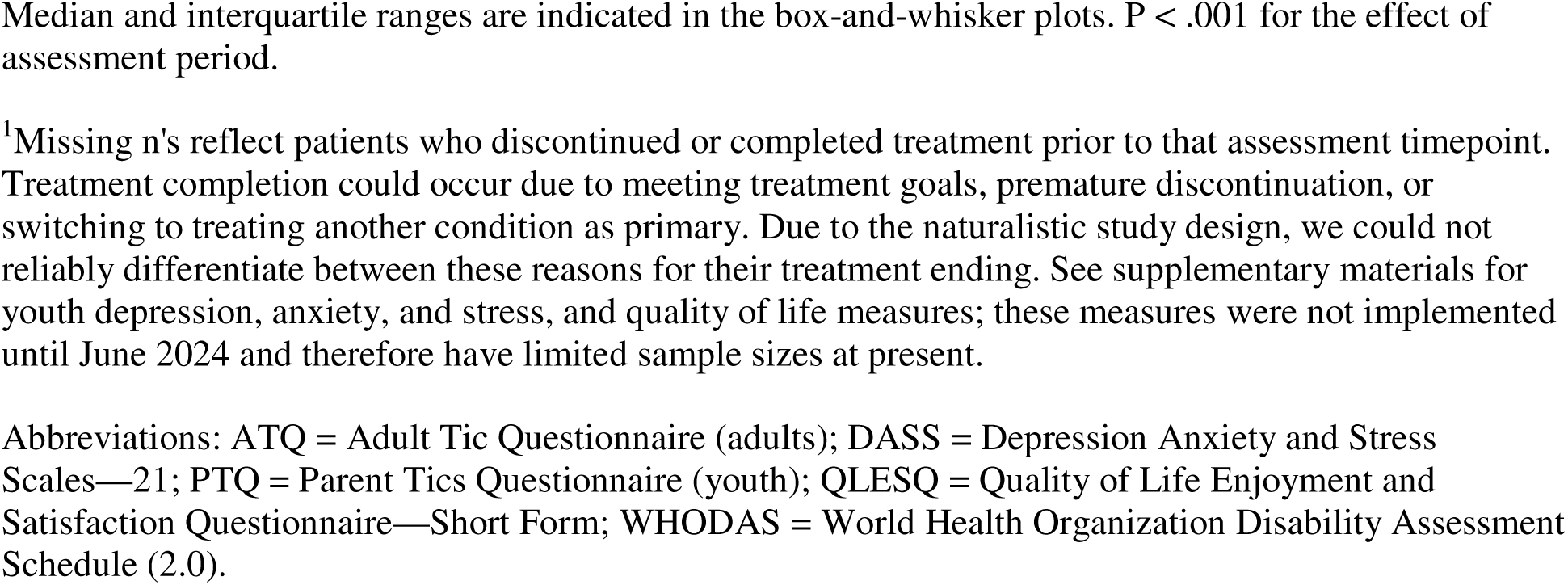
Clinical assessments by treatment time point.

|  | **Youth** |  |  | **Adult** |  |  |
| --- | --- | --- | --- | --- | --- | --- |
| Outcome scale and assessment time point | Valid, n | Missing<sup>1</sup>, n | Mean (SD) | Valid, n | Missing<sup>1</sup>, n | Mean (SD) |
| **PTQ/ATQ** |  |  |  |  |  |  |
| Baseline | 114 | 0 | 28.8 (21.0) | 53 | 0 | 39.3 (28.8) |
| Session 7 | 101 | 13 | 22.6 (18.5) | 45 | 8 | 28.0 (22.0) |
| Session 14 | 61 | 53 | 18.6 (21.3) | 31 | 22 | 29.8 (25.7) |
| **DASS depression** |  |  |  |  |  |  |
| Baseline | — | — | — | 50 | 3 | 9.9 (10.3) |
| Session 7 | — | — | — | 42 | 11 | 8.5 (8.2) |
| Session 14 | — | — | — | 29 | 24 | 9.4 (10.2) |
| **DASS anxiety** |  |  |  |  |  |  |
| Baseline | — | — | — | 50 | 3 | 6.8 (7.9) |
| Session 7 | — | — | — | 42 | 11 | 5.9 (5.9) |
| Session 14 | — | — | — | 29 | 24 | 6.1 (7.2) |
| **DASS stress** |  |  |  |  |  |  |
| Baseline | — | — | — | 50 | 3 | 16.0 (10.5) |
| Session 7 | — | — | — | 42 | 11 | 12.7 (8.4) |
| Session 14 | — | — | — | 29 | 24 | 11.5 (9.6) |
| **Q-LES-Q** |  |  |  |  |  |  |
| Baseline | — | — | — | 50 | 3 | 62.9 (16.4) |
| Session 7 | — | — | — | 43 | 10 | 66.4 (14.0) |
| Session 14 | — | — | — | 30 | 23 | 64.9 (17.3) |
| **WHODAS** |  |  |  |  |  |  |
| Baseline | — | — | — | 49 | 4 | 21.1 (9.2) |
| Session 7 | — | — | — | 42 | 11 | 19.7 (7.0) |
| Session 14 | — | — | — | 21 | 32 | 20.2 (6.5) |

Median and interquartile ranges are indicated in the box-and-whisker plots. $P < .001$ for the effect of assessment period.
<sup>1</sup>Missing n's reflect patients who discontinued or completed treatment prior to that assessment timepoint. Treatment completion could occur due to meeting treatment goals, premature discontinuation, or switching to treating another condition as primary. Due to the naturalistic study design, we could not reliably differentiate between these reasons for their treatment ending. See supplementary materials for youth depression, anxiety, and stress, and quality of life measures; these measures were not implemented until June 2024 and therefore have limited sample sizes at present.
Abbreviations: ATQ = Adult Tic Questionnaire (adults); DASS = Depression Anxiety and Stress Scales—21; PTQ = Parent Tics Questionnaire (youth); QLESQ = Quality of Life Enjoyment and Satisfaction Questionnaire—Short Form; WHODAS = World Health Organization Disability Assessment Schedule (2.0).

### 3.7 Engagement and Satisfaction

Patients demonstrated high engagement with the virtual platform, with 94.6% using in-app messaging and a median satisfaction rating of 5.0 [IQR: 5-5] on a 5-point scale (mean: 4.78, SD: 0.57), with no significant differences between age groups (see Supplementary Materials for detailed engagement metrics).

## 4. Discussion

This is the first large-scale retrospective observational longitudinal analysis of clinical outcomes of a naturalistic sample of individuals with tic disorders treated with therapist-delivered video therapy (HRT) with digital support. Our findings demonstrate that this approach is effective in reducing tic symptoms across the lifespan. Specifically, patients showed a median 39.0% reduction in tic severity at session 14, with 54.3% achieving at least a 35% reduction in symptoms and 35.9% achieving a 55% or greater reduction. Youth showed a numerically (slightly) stronger treatment response (median 40.9% reduction) compared to adults (median 38.1% reduction), although this difference was not statistically significant. Improvements continued to strengthen until the final session, with the combined sample showing a median reduction of 44.4% from baseline at final session completion, suggesting potential continued benefits of extended therapist-delivered video therapy HRT with digital support. This aligns with other studies showing sustained improvements in tic severity and related impairments at follow-ups ranging from several months to years^41,42^. Additionally, treatment led to significant improvements in commonly co-occurring symptoms of depression, anxiety, and stress, as well as enhancements in quality of life and functioning. These broader improvements are noteworthy given that tic disorders can significantly impact school performance, occupational functioning, and social relationships^43–45^.

These findings have important implications given that tics typically onset in childhood and frequently persist into adulthood if untreated. There are often substantial delays between symptom onset and treatment access, with studies showing delays due to multiple factors including misdiagnosis, insidious symptom development, lack of access to trained therapists, and comorbid conditions that can complicate the clinical picture^46–49^. Without appropriate intervention, tic disorders can significantly impair quality of life across multiple domains, affecting physical, social, familial, academic, and psychological functioning^9–12^.

Tic disorders impose substantial economic and societal burdens that extend far beyond direct medical costs. Children with tic disorders require care from multiple healthcare professionals, leading to high healthcare utilization and inefficient service use due to inadequate care pathways and lack of specialized providers^22^. Families experience significant indirect costs through school absenteeism as children miss school to access care, while both patients and families report reduced quality of life and increased stress, particularly when comorbid conditions are present^11,50^. The burden extends beyond families to schools and communities, with increased need for support and accommodations ^51^. Evidence suggests that implementing accessible, evidence-based online behavioral therapies could generate substantial healthcare cost savings, potentially up to £1 million for systems like the U.K.’s National Health Service (NHS), by reducing unnecessary specialist contacts and improving care efficiency^22^. Our therapist-delivered video therapy HRT with digital support directly addresses these critical access barriers, offering a scalable solution that could reduce downstream healthcare costs while improving population health outcomes.

Our findings align with previous research on the effectiveness of HRT and CBIT for tic disorders, while extending this evidence to a therapist-delivered video therapy format.. Meta-analyses of in-person HRT have typically found small to medium effect sizes (SMD of-0.43) in reducing tic symptoms^16^, while CBIT has demonstrated moderate to large effect sizes across studies^52,53^. Our effect sizes for this approach (g=0.47 overall at session 14, increasing to g=0.57 at the final session) are comparable to these previously reported ranges for in-person HRT, suggesting that HRT delivered in a therapist-delivered video therapy format can achieve similar effectiveness to traditional in-person delivery.

Previous research has reported results for videoconference and VoIP-delivered behavioral interventions, other than HRT, for tic disorders. Studies of CBIT delivered via videoconferencing have reported a range of effect sizes, including large effects for youth (Cohen’s d=1.31) and medium-to-large effects for adults (Cohen’s d=0.66) in a single-arm trial^24^. Smaller randomized controlled trials have shown more modest effect sizes (partial η²=0.15-0.26) for VoIP-delivered behavior therapy compared to waitlist controls^23^. Effect sizes for our approach (Hedges’ g=0.47 overall at session 14, increasing to g=0.57 at final session) are generally consistent with the Cohen’s d values reported in previous studies. However, direct comparisons of these effect sizes with those from our sample should be made with caution, both because Cohen’s d and Hedge’s g are closely related but distinct measures of effect size (particularly when accounting for differences in sample sizes) and the previous studies were clinical trials rather than observational results.

Separately, self-guided and self-guided/therapist-supported internet-based behavioral interventions have also demonstrated promising effectiveness for tic disorders^54–56^. These approaches typically deliver structured therapeutic content online with minimal therapist support. The Online Remote Behavioural Intervention for Tics (ORBIT) trial, a large multicenter randomized controlled trial (n=224) found that therapist-supported online Exposure and Response Prevention (ERP) produced modest, but durable benefits compared to psychoeducation, with between-group effect sizes of-0.31 at 3 months and - 0.27 at 18 months^54,57^. Similar results emerged from Andrén and colleagues’, which began with a pilot study suggesting superior efficacy for internet-delivered ERP (d=1.12) over HRT (d=0.50) and was followed by a full RCT (n=221) demonstrating ERP’s higher treatment response rates (47% vs. 29%) compared to psychoeducation^55,58,59^. Research on internet-based guided self-help CBIT has shown even larger effects, with initial improvements (d=0.91) strengthening at 6-month follow-up (d=2.25), even among patients with comorbid conditions^60,61^. Our effect sizes are comparable to these findings, although direct comparisons are complicated by differences in delivery format (therapist-delivered video therapy with digital support versus self-guided with minimal support), specific intervention components, and the aforementioned differences between Cohen’s d and Hedge’s g. Nevertheless, the promising outcomes observed in both our therapist-delivered video therapy format and these internet-based programs underscore the potential of remote behavioral interventions for individuals with tic disorders.

Patients completed a median of 15 sessions over approximately 20.9 weeks (for sessions 1-14), with 55.1% completing ≥14 primary treatment sessions. This timeframe is consistent with other HRT protocols that typically span 10-16 weeks across various delivery formats ^55,62–64^. Most controlled research on HRT implements 8-14 weekly sessions, with examples including eight weekly sessions in adults ^65^ and children ^19^, nine sessions in mixed group and individual formats ^63^, and 14 sessions for adults with Tourette’s disorder ^66,67^. Some studies have used more intensive protocols, such as 16 sessions for children and adolescents ^62^, while others have demonstrated efficacy with abbreviated approaches, including a modified 4-session program ^41^. The ORBIT trial similarly utilized a 10-week intervention period for their internet-delivered behavioral therapy ^48,57^. Importantly, our analysis of the final session (median total of 15 sessions over 30.9 weeks) suggests that flexible treatment duration may optimize outcomes, with 92% of patients who completed ≥14 sessions engaging in additional maintenance therapy. The flexible option for an extended timeframe may have contributed to the robust outcomes observed, while still maintaining efficiency compared to traditional in-person treatments that can sometimes extend to several months of weekly sessions ^18^.

The median session frequency was 0.6 sessions per week (approximately 2-3 sessions per month), which is lower than the weekly frequency typically used in controlled trials. The naturalistic delivery pattern of approximately 2-3 sessions per month likely reflects real-world constraints that are rarely present in highly controlled clinical trial environments. For example, some of our patients had already been in therapy at NOCD for other conditions before starting tic treatment. For patients with prior treatment for conditions, for example OCD, their earlier exposure to behavioral principles may have facilitated more efficient tics treatment, as HRT shares conceptual foundations with other behavioral interventions. Additionally, practical constraints in real-world settings often make weekly sessions challenging (e.g., school or work commitments, caregiver schedules for youth patients, and competing healthcare appointments). Yet, these logistical challenges tend to be more difficult for in-person treatment, while our therapist-delivered video therapy format eliminated geographical barriers and travel burden, potentially enhancing accessibility to specialized treatment for patients in areas with limited access to HRT specialists.

Several unique features of NOCD’s integrated treatment approach may have contributed to the observed outcomes. The video therapy format allowed patients to engage in treatment within their natural environments, potentially enhancing ecological validity and generalization of skills. This advantage is particularly relevant for tic disorders, as interventions addressing symptoms in natural contexts can better support improved school performance and social functioning^43,68^. Importantly, this approach goes beyond traditional telehealth by integrating multiple digital support components that work synergistically with the video therapy sessions.Patients had access to between-session support through in-app messaging with their therapist, a mobile app journal tool for target behavior monitoring, and a moderated online community. This integrated combination of live video therapy with digital support tools created a comprehensive therapeutic ecosystem that extended engagement beyond scheduled sessions, enabling continuous symptom tracking and peer connection while maintaining ongoing therapist communication between sessions. Previous studies have found telehealth to be both feasible and acceptable, with high patient satisfaction ratings and strong therapeutic alliances^24^. These positive patient experiences likely contribute to treatment engagement and may represent a preference for the convenience and accessibility of virtual therapy over traditional in-person formats, particularly for individuals in underserved areas or those with transportation limitations.

Our analysis revealed interesting patterns across demographic and severity subgroups. Treatment outcomes showed some variation across age groups, with youth demonstrating slightly stronger median improvement (40.9% at session 14) compared to adults (38.1% at session 14), although both groups achieved clinically meaningful reductions. Notably, adults showed the steepest improvements from baseline to session 7 (28.8% reduction), with some leveling off at session 14 (24.2% reduction). This pattern may reflect both the typical trajectory of early gains in behavioral interventions and selective attrition, where patients achieving good early responses may discontinue treatment while others continue. This suggests that therapist-delivered video HRT with digital support can be effectively adapted for patients at different developmental stages. Our baseline severity analysis also revealed important moderating effects that differed between age groups. Among youth, baseline severity was positively associated with absolute symptom reduction at both session 7 (r=.50, p<.001) and session 14 (r=.51, p<.001), indicating that youth with more severe symptoms showed greater absolute improvement while achieving comparable percentage improvements to those with milder symptoms. Adults showed a different pattern, with baseline severity positively associated with absolute reduction at session 7 (r=.47, p=.001), but this relationship disappeared by session 14 (r=-.03, p=.88) and trended negative for percent reduction (r=-.30, p=.10). These differential patterns suggest that youth with higher baseline severity may be particularly good candidates for this approach, while adult outcomes appear less dependent on initial symptom severity over the course of treatment.

Several limitations warrant discussion. The observational design without a control group or randomization precludes causal inferences about treatment effects, although symptom improvements are similar to those in previous controlled trials of HRT. Additionally, while we used validated measures (PTQ and ATQ) to assess tic symptoms, these self-report and parent-report measures may be subject to reporter bias. While established clinical response cutoffs have been validated for the PTQ in youth populations^40^ comparable validated thresholds have not been established for the ATQ in adults. To maintain consistency and interpretability across age groups, we applied uniform percentage-based response criteria rather than using the validated dual criteria for youth, which may limit direct comparison with previous PTQ-based studies. It is important to acknowledge the considerable attrition from our initial pool of 387 patients to our final analyzed sample of 167 (43.2%). We implemented specific inclusion criteria requiring baseline assessment completion (sessions 1-2), at least one follow-up assessment, and minimum five tic-focused sessions to ensure meaningful treatment engagement. Our analysis used last-observation-carried-forward (LOCF) methods for missing assessments, providing conservative estimates that do not overestimate treatment effects. However, our findings may not generalize to patients who discontinued early or had minimal treatment engagement. Another limitation is the low proportion of patients with severe symptoms in our sample, which limits generalizability to severe tic presentations and highlights the need for future research across the full spectrum of symptom severity. Further, without formal treatment fidelity assessments, we cannot definitively determine therapist adherence to the HRT protocol, despite standardized training and supervision. Future exploration and comparison of emerging telehealth strategies and treatments for tics, such as gamified interventions^69^ and online mindfulness-based approaches ^70^, could help evaluate potential expanded options for effective telehealth delivery of tic disorder treatments.

In conclusion, this analysis demonstrates that therapist-delivered video HRT with digital support is effective in reducing tic severity and improving related symptoms across the lifespan in a real-world setting. With median improvements of 39.0% at session 14 and continued benefits at the final session (44.4% median improvement), the treatment effects suggest this approach may help address barriers to accessing evidence-based care for tic disorders. By leveraging technology to deliver specialized behavioral interventions, we can potentially bridge the substantial gap between symptom onset and effective treatment that many individuals with tic disorders currently experience.

## Data Availability

The data sets analyzed during this study are proprietary business assets of NOCD Inc., and are not publicly available. The data may be available from the corresponding author on reasonable request with appropriate data use agreements in place.

## Acknowledgements

The authors thank the NOCD therapists and Member Advocates for their help in facilitating treatment and care experiences.

## Relevant Financial Relationships

JDF, CCB, PBM, NRF, MN, LT, SMS, AR, and NL report personal fees from NOCD Inc.

