## SupplementalFiles for "Habit Reversal Training for Tic Disorders: Clinical Outcomes from a Large Sample of Youth and Adults Treated with Therapist-delivered Video Therapy"

**Supplementary Materials**

**Method**

*Sample*

**Supplementary Figure 1**. Application of inclusion criteria to derive the final analysis sample.

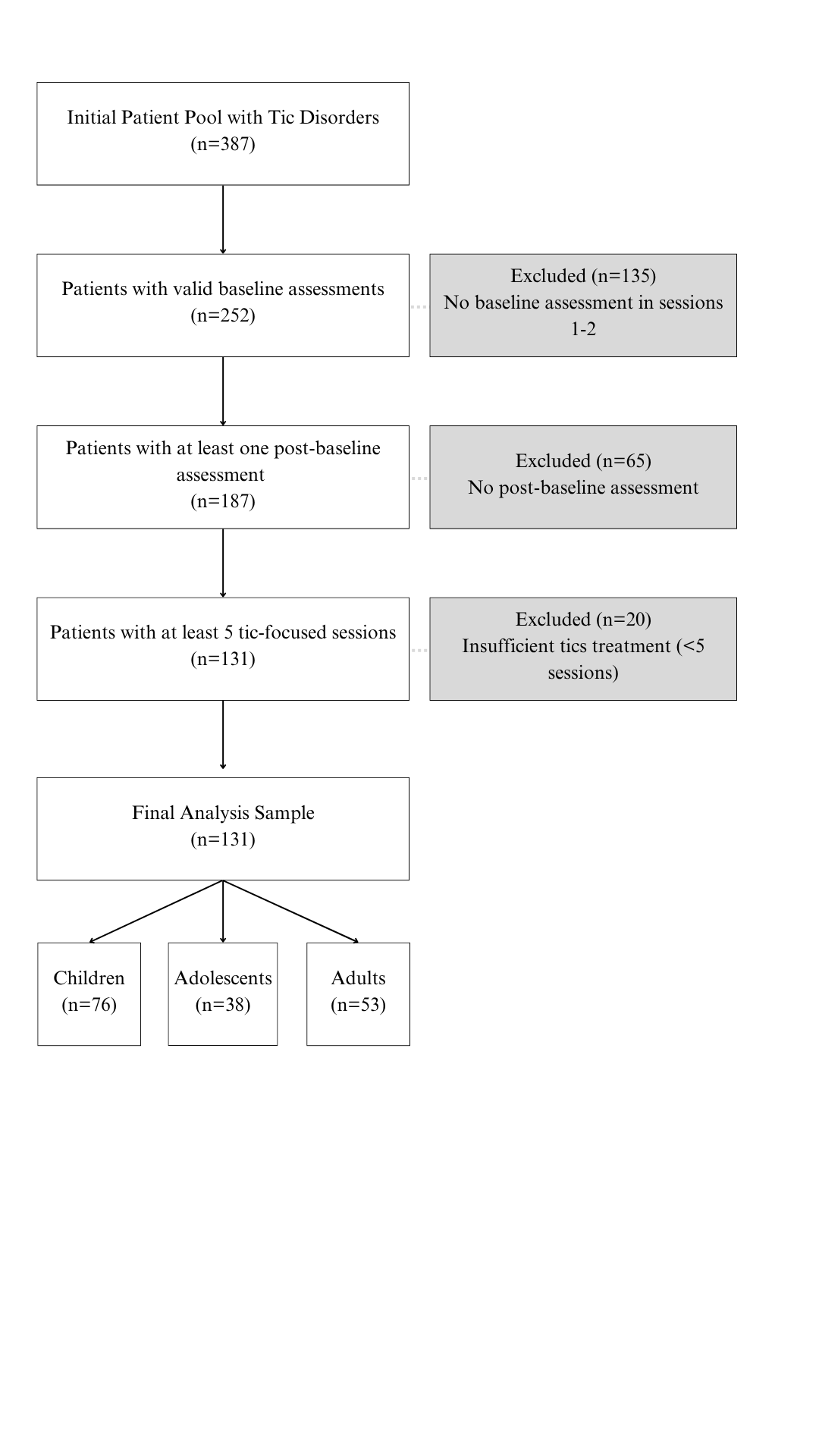

*Initial Evaluation and Clinical Assessments*

Patients entered treatment through NOCD's standard clinical pathway, either through self-referral or referral from insurance providers or medical professionals. Initial diagnostic evaluations were conducted by NOCD-trained therapists over two sessions, comprising a comprehensive biopsychosocial assessment and a semi-structured diagnostic interview using the Diagnostic Interview for Anxiety, Mood, and Obsessive-Compulsive and Related Neuropsychiatric Disorders (DIAMOND^1,2^) Patients who met DSM-5 criteria for persistent (chronic) motor or vocal tic disorder, or Tourette syndrome, were eligible for treatment. In cases in which a patient had a tic disorder comorbid with another disorder that NOCD treats, such as OCD, obsessive-compulsive related disorders, or comorbid OCD and posttraumatic stress disorder, the therapist and patient or parent/caregiver would decide which disorder should be treated first and then, typically, treat the other disorder second. NOCD typically serves individuals aged 5 and older. Additional referrals were made as needed for concurrent psychiatric (other than those treated by NOCD) or substance use concerns that might interfere with HRT treatment.

*Therapist Qualifications and Training*

Therapists completed an intensive 12 week training at NOCD on the treatment of OCD and obsessive-compulsive related disorders, as well as commonly co-occurring conditions. Training specific to treatment of tics was delivered in a hybrid model consisting of both live and asynchronous didactic learning with role play demonstrations followed by a post-test to ascertain knowledge and skills accrued. Therapists attended consultation groups to receive ongoing guidance on treatment delivery, as well as to ask case-specific questions.

*Secondary Outcome Measures*

1. Depression, Anxiety, and Stress Scale (DASS-21^3^): 21-item self-report measure with three 7-item subscales assessing depression, anxiety, and stress in adults. Items are rated from 0 ("Did not apply to me at all") to 3 ("Applied to me very much"), with subscale scores multiplied by 2 to yield final scores ranging from 0-42, where higher scores indicate greater symptom severity.
2. Depression, Anxiety, and Stress Scale-Youth (DASS-Y^4^): 21-item version adapted for children and adolescents, maintaining the same three-factor structure, rating scale, and scoring methods as the adult version, with demonstrated psychometric properties for ages 7-18.
3. Quality of Life Enjoyment and Satisfaction Questionnaire-Short Form (Q-LES-Q-SF^5^): 16-item self-report measure for adults with the first 14 items scored on a 5-point scale and summed to yield a total percentage score ranging from 0-100, where higher scores indicate better quality of life.
4. Pediatric Quality of Life Enjoyment and Satisfaction Questionnaire (PQ-LES-Q^6^): 15-item adaptation for children and adolescents aged 6-17, using the same 5-point rating scale and percentage scoring system as the adult version.
5. World Health Organization Disability Assessment Schedule 2.0 (WHODAS 2.0^7^): 12 item self-report measure assessing disability and functioning related to medical and/or mental health conditions. Items are scored from 0 (no difficulty) to 4 (extreme difficulty), with total scores ranging from 0-48. Higher scores indicate greater impairment.

*Data Analysis*

Missing data analyses were conducted to determine whether data were Missing Completely At Random (MCAR) by comparing baseline characteristics between participants with early discontinuation (≤7 sessions, n=29) versus sustained engagement (8+ sessions, n=138). The 7-session threshold was chosen because 8 sessions represents the minimum standard HRT/CBIT protocol in the literature^8^. Treatment satisfaction and treatment engagement metrics (messaging, app opens, visits) were calculated using cumulative engagement data from each patient's final treatment session and the most recent available satisfaction rating, analyzing message frequency and app usage patterns across the full treatment duration. Exploratory analyses also examined the impact of baseline severity and treatment parameters (session frequency, duration) on treatment outcomes. Secondary measures assessed youth depression, anxiety, and stress (DASS-Youth DASS-Y[^4^) and quality of life (Pediatric Quality of Life Enjoyment and Satisfaction Questionnaire PQ-LES-Q[^6^]. Secondary outcomes were analyzed using linear mixed models with treatment session (baseline, session 7, session 14) as a fixed factor, patient as a random factor, and respective outcome scores (depression, anxiety, stress, quality of life) as dependent variables.

**Results**

*Treatment Delivery and Engagement*

The distribution of treatment length showed considerable variation: 17.4% completed <8 sessions, 31.7% completed 8-14 sessions, 25.7% completed 15-24 sessions, and 25.1% completed 25+ sessions, with total sessions ranging from 5 to 72 across all patients.

*Primary Treatment Phase Completion (Sessions 1-14).* Completion rates were similar between adults (58.5%, n=53) and youth (53.5%, n=114, p=0.663). Adults and youth took a median of 20.9 weeks to complete the primary phase, though adults showed greater variability [IQR: 12.9-33.4] compared to youth [IQR: 14.9-26.9].

*Maintenance Phase Engagement*. Among patients who completed ≥14 sessions, 92.4% (85/92) engaged in additional maintenance therapy, with only 7 patients stopping at exactly session 14. These 85 patients completed a median total of 24 sessions [IQR: 17-33], including 10 additional maintenance sessions [IQR: 3-19] over a median duration of 21.9 weeks [IQR: 7.9-42.9]. Adults in the maintenance phase (n=28) completed a median of 27 total sessions [IQR: 19.5-33.25] compared to 23 sessions [IQR: 17-33] for youth (n=57), though this difference was not statistically significant (p=0.51).

*Missing Data (MCAR)*

For adults (n=53), no significant differences were found between early discontinuation (n=11) and sustained engagement (n=42) groups for age at therapy start (t(12.93)=0.62, p=.55), gender distribution (χ²(2)=4.17, p=.125), or baseline tic severity (t(11.42)=0.57, p=.58). Adult secondary measures showed no significant differences for DASS anxiety, depression, stress, or total scores (all p>.05, n=50), WHODAS functional impairment (p=.84, n=49), or Q-LES-Q quality of life (p=.38, n=50).

For youth (n=114), a significant difference was found for sex distribution between early discontinuation (n=18) and sustained engagement (n=96) groups (χ²(2)=14.25, p<.001. Early discontinuation had 4 female (22.2%), 9 male (50.0%), and 5 unknown (27.8%) participants, while sustained engagement had 34 female (35.4%), 59 male (61.5%), and 3 unknown (3.1%) participants. The pattern suggests higher rates of missing gender data among early treatment discontinuers. No significant differences were found for age at therapy start (t(21.67)=-0.56, p=.58) or baseline tic severity (t(27.48)=-0.23, p=.82). Youth secondary measures could not be reliably assessed due to limited sample sizes.

*Treatment Engagement and Satisfaction*

Patients demonstrated active engagement with the app. They opened the app a median of 62 times [IQR: 36.5-111.5] throughout treatment, with considerable variation (mean: 90.4, SD: 89.5). The vast majority of patients (94.6%) engaged in in-app messaging, with a median of 40.5 messages [IQR: 23-83] exchanged among those who used this feature.

App usage patterns were similar between youth and adults. Adult patients opened the app a median of 69 times [IQR: 35-116] compared to youth patients (which included both youth and parent messaging) (median: 59.5 [IQR: 37-108]), with no statistically significant difference (p=0.425). Messaging engagement was high across both groups, with 90.6% of adults and 96.5% of youth exchanging messages. Among those who used messaging, adults exchanged slightly more messages (median: 44.5 [IQR: 26.5-84.25]) than youth (median: 36.5 [IQR: 20-80]), though this difference was not statistically significant (p=0.226).

Treatment satisfaction ratings were available for 65.3% of patients (n=109), with a median rating of 5.0 [IQR: 5-5] on a 5-point scale (mean: 4.78, SD: 0.57). Satisfaction ratings were similarly high between youth (n=78, median: 5.0 [IQR: 5-5], mean: 4.80) and adults (n=31, median: 5.0 [IQR: 5-5], mean: 4.73), with no significant difference between age groups (p=0.648).

*Treatment parameter effects*

A three-way interaction analysis (timepoint × treatment duration × age group) found no significant differences in how duration influences treatment trajectories between youth and adults (F(2, 176)=1.55, p=.216). Among youth, the interaction between timepoint and duration was not significant (F(2, 118)=1.60, p=.206). Treatment duration showed no meaningful relationship with either absolute reduction (r(59)=.13, p=.336), or percent reduction in tic symptoms (r(59)=.12, p=.340), suggesting that youth respond similarly to treatment regardless of its duration. Adults similarly showed no significant timepoint × duration interaction (F(2, 58)=0.49, p=.615). However, unlike youth, adults exhibited a trend toward better percentage improvement with longer treatment (r(29)=.33, p=.069), although this relationship didn't reach statistical significance. No association was found between duration and absolute symptom reduction (r(29)=.10, p=.603).

*Youth Depression, Anxiety, Stress, and Quality of Life*

Youth secondary measures were implemented beginning in June 2024, resulting in limited sample sizes (see Supplementary Table 1). Linear mixed model analysis showed that stress had the most consistent improvement pattern (p=.02), with effect sizes ranging from g=0.23 to g=0.40 across timepoints. Depression and anxiety showed mixed patterns, with some individual timepoints showing promise but overall inconsistent effects. Youth quality of life on the PQLESQ showed minimal change across all timepoints, with effect sizes near zero and no significant overall time effect (p=.77). These findings suggest that stress symptoms were most responsive to treatment among youth, while depression, anxiety, and quality of life showed more limited improvements. Results should be interpreted cautiously given the smaller sample sizes (see Supplementary Table 1 for complete results).

**Supplementary Table 1.** Youth Secondary Outcome Measures

| **Measure** | **Timepoint** | **N** | **Mean ± SD** | **Effect Size (95% CI)** | **Median % Change (IQR)** | **p-value** |
| --- | --- | --- | --- | --- | --- | --- |
| DASS-Y Depression | Baseline | 30 | 1.93 ± 3.47 | -- | -- | .19 |
|  | Session 7 | 28 | 1.16 ± 2.21 | 0.28 [-0.03, 0.59] | 64.3 [0.0, 100.0] |  |
|  | Session 14 | 40 | 0.93 ± 1.75 | 0.39 [-0.23, 1.01] | 100.0 [-25.0, 100.0] |  |
|  | Final Session | 27 | 1.47 ± 2.66 | 0.14 [-0.17, 0.44] | -12.5 [-∞*, 100.0] |  |
| DASS-Y Anxiety | Baseline | 30 | 2.43 ± 4.22 | -- | -- | .29 |
|  | Session 7 | 28 | 2.16 ± 4.18 | 0.02 [-0.18, 0.22] | 0.0 [-100.0, 0.0] |  |
|  | Session 14 | 40 | 0.87 ± 1.85 | 0.26 [-0.01, 0.54] | 85.7 [27.3, 100.0] |  |
|  | Final Session | 27 | 1.69 ± 3.38 | 0.07 [-0.14, 0.28] | 0.0 [-25.0, 60.7] |  |
| DASS-Y Stress | Baseline | 30 | 8.47 ± 4.99 | -- | -- | .02 |
|  | Session 7 | 28 | 6.94 ± 4.18 | 0.23 [0.01, 0.45] | 0.0 [0.0, 25.0] |  |
|  | Session 14 | 46 | 6.20 ± 4.00 | 0.40 [-0.06, 0.86] | 31.7 [0.0, 52.9] |  |
|  | Final Session | 27 | 6.78 ± 4.12 | 0.27 [0.02, 0.51] | 0.0 [0.0, 34.3] |  |
| PQLESQ | Baseline | 35 | 57.51 ± 6.90 | -- | -- | .77 |
|  | Session 7 | 32 | 58.05 ± 5.62 | 0.18 [0.01, 0.35] | 0.0 [0.0, 3.9] |  |
|  | Session 14 | 15 | 57.30 ± 6.09 | 0.00 [-0.46, 0.46] | -1.7 [-4.7, 8.9] |  |
|  | Final Session | 32 | 57.05 ± 7.09 | -0.02 [-0.28, 0.24] | 0.0 [-5.0, 5.5] |  |

*Effect sizes are Hedges' g. P-values are from linear mixed models testing the overall time effect (whether scores changed significantly across baseline, session 7, session 14, and final session timepoints). *∞ indicates percentage change calculations involving baseline scores of zero*

Abbreviations: DASSY = Depression, Anxiety, and Stress Scale-Youth; PQLESQ = Pediatric Quality of Life Enjoyment and Satisfaction Questionnaire.
